# Granular Insights:A Wastewater-Based Machine Learning Approach for Localized COVID-19 Hospitalization Forecasting

**DOI:** 10.1101/2025.06.25.25330294

**Authors:** Nusrat Tabassum, Mohammad Mihrab Chowdhury, Christopher S McMahan, Stella Self, Mirza Isanovic, Karlen Correa-Velez, Sarah C. Sellers, R. Sean Norman, Lior Rennert

## Abstract

Wastewater based epidemiology (WBE) is a valuable tool for monitoring emerging disease trends in a community. Specifically, early predictions of hospitalization in a community can help reduce the strain on healthcare services and facilitate better planning and preparation. This study examines the use of SARS-CoV-2 RNA concentrations in wastewater to predict COVID-19 hospitalizations in South Carolina. We analyzed SARS-CoV-2 RNA concentration collected from six wastewater treatment plants (WWTPs) across South Carolina from April 19, 2020 to February 2, 2021 to predict COVID-19-related hospitalizations across WWTPs and 43 associated ZIP codes. Poisson regression and random forest models were utilized to forecast 7-day, 14-day, and 21-day ahead COVID-19 hospitalizations. Model performance was validated against statewide hospitalization claims data. Model accuracy was strongest for 14-day ahead prediction, with the random forest models achieving a median percentage agreement (PA) of 91.16% (IQR = 86.49–91.84%) across WWTPs and 78.12% (IQR = 67.99–84.53%) across ZIP codes. These findings demonstrate that WBE offers a robust and timely approach for predicting hospitalizations at fine geographic scales. This modeling framework can be adapted to other infectious diseases to enhance surveillance and response efforts.

## 1. Introduction

Wastewater surveillance has become an effective tool in public health, allowing the monitoring of community health through the analysis of viral and microbial biomarkers found in sewage. Wastewater-based epidemiology (WBE) provides population-level insights into circulating diseases and health conditions by detecting biomarkers, pathogens, and chemicals in wastewater (Petersen and Lupton, 2000; McCarthy and Shugart, 1990; Sims and Kasprzyk-Hordern, 2020; Zuccato et al., 2005). The first documented link between disease and wastewater was in the 1854 Broad Street cholera outbreak in London, traced by Dr John Snow, widely regarded as the father of epidemiology (Wilson, 1933; Tulchinsky, 2018; Cameron and Jones, 1983; Buechner et al., 2004). In the 1940s, wastewater-based epidemiology (WBE) was utilized in the U.S. to track poliovirus outbreaks (Kelly et al., 1957; Metcalf et al., 1995). Eventually, WBE played a key role in global polio eradication despite the lack of standardized methods until the 1990s (Brouwer et al., 2018; Asghar et al., 2014).The introduction of PCR in the 1990s marked a turning point, establishing the gold standard for wastewater pathogen detection (Toze, 1999; Erlich et al., 1991). During the COVID-19 pandemic, WBE emerged as a key early warning system for disease outbreaks (Larsen et al., 2020; Hillary et al., 2020; Maida et al., 2022; Haramoto et al., 2020; Ahmed et al., 2020). The pandemic has profoundly disrupted global public health systems, exposing the vulnerabilities in disease monitoring and resource allocation (Haldane et al., 2021). It has caused over 700 million infections and over 7 million deaths worldwide (Dec., 2019-August, 2024) (Feldscher, 2024; Feehan and Apostolopoulos, 2021). In U.S., COVID-19 was a major contributor to mortality, ranking as the third leading cause of death in 2020 and 2021, and the fourth in 2022 (Ahmad et al., 2023; Murphy et al., 2021; Ahmad et al., 2022). As of April 26, 2023, over 104 million COVID-19 cases, 6 million hospitalizations, and 1.1 million deaths were reported in the U.S. (Silk, 2023). The large volume of cases and hospitalizations has heavily strained healthcare systems and public health infrastructure. Alongside this, the COVID-19 pandemic in the U.S. led to an estimated direct medical cost of $163.4 billion (Bartsch et al., 2020). Indirect costs, including absenteeism and productivity losses, averaged $671.4 per patient (Faramarzi et al., 2021). Globally, the economic burden ranged from $77 billion to $2.7 trillion (Forsythe et al., 2020; Miles et al., 2021; Huo et al., 2021; Zala et al., 2020; Zhao et al., 2021; Ebigbo et al., 2021). This dual burden on healthcare systems and the economy highlights the urgent need for timely, accurate, and innovative tools to guide early public health interventions and preparation.

Traditional methods for estimating infectious disease trends rely heavily on case counts, seroprevalence surveys, or hospitalizations. While effective at the state or national level, these methods often fail to provide timely, localized insights (Lipsitch et al., 2011; Metcalf et al., 2016; Pullano et al., 2021). Delays in data reporting, limited geographic specificity, testing availability, case definitions, and reporting standards (tested or confirmed cases) often bias estimates, hindering community-level decisions (Hart and Halden, 2020; Islam et al., 2021). Additionally, symptomatic cases dominate traditional surveillance, which significantly underestimates the true burden of disease (Fraser et al., 2009; Oran and Topol, 2020). Such limitations make it hard to identify disease trends in small or low-access areas. Wastewater surveillance helps address these challenges by quantifying SARS-CoV-2 RNA copies in sewage, capturing data from both symptomatic and asymptomatic individuals (Hart and Halden, 2020; Huisman et al., 2022). It is not biased by healthcare access and offers a cost-effective, scalable solution for early outbreak detection and trend forecasting (Bibby et al., 2021; D’Aoust et al., 2021). This makes WBE a valuable resource for real-time monitoring of infectious diseases in granular regions.

Early studies have demonstrated that WBE can detect SARS-CoV-2 RNA in wastewater with a lead time of one to two weeks compared to clinical testing methods (Galani et al., 2022; La Rosa et al., 2020). Long-term surveillance shows that higher sampling frequency improves detection (Feng et al., 2021; Graham et al., 2020). Machine learning (ML) models—including random forests and artificial neural networks (ANNs)—have been used to forecast COVID-19 trends from wastewater data (Li et al., 2023; Jiang et al., 2022; Kanchan et al., 2024). However, despite recent advancements, the application of WBE often lacks the geographic granularity needed for detailed public health planning. WBE data are typically aggregated at the wastewater treatment plant (WWTP) level, limiting the ability to detect localized hospitalization trends. ZIP code-level data are essential for identifying patterns that enable targeted interventions(Rennert et al., 2024; Hossain et al., 2025; Gezer et al., 2024b). Specifically, hospitalizations, which heavily burden healthcare systems, are often overlooked at this finer resolution. Previous research highlights the importance of geographic granularity in effective pandemic response and resource planning. For instance, ZIP code-level analysis enables the strategic deployment of mobile health clinics (MHCs) and other resources to underserved areas(Rennert et al., 2024; Gezer et al., 2024a) . This detail helps identify communities with high disease burdens or limited access to healthcare, allowing for targeted interventions such as localized testing, emergency response, and real-time resource allocation.

Recognizing these gaps, our study presents a wastewater-based modelling framework to predict COVID-19 hospitalizations at both the ZIP code and WWTPs, supporting data-driven public health decision-making. Leveraging RNA copy data from WWTPs in South Carolina (SC), the framework employs machine learning models to forecast hospitalizations. Extending the work of Swift et al. (2023), on estimating infection from RNA at WWTP, we incorporate socio-demographic characteristics and granular region (ZIP code) to capture community-level dynamics better. The resulting framework offers public health agencies a reliable tool for allocating resources and responding to localized disease burdens. Beyond COVID-19, this modelling approach is adaptable to other infectious diseases, enhancing preparedness and targeted intervention across diverse public health spectrum.

## 2. Materials and Methods

### 2.1. Study Setting

A statewide wastewater surveillance initiative in SC monitored SARS-CoV-2 RNA concentrations across geographically diverse WWTPs (Swift et al., 2023). This study focused on a total of six WWTPs situated in Richland, Lexington, York, and Charleston. These areas collectively served approximately a 1.4 million individuals, representing nearly 25.9% of the state’s population. These counties reflect South Carolina’s socio-demographic and geographic diversity, making them valuable for this study. Richland (429,863 people) and Charleston (419,279 people) are urban centers with major healthcare facilities, while Lexington (300,906 people) and York (289,620 people) include suburban and rural communities, capturing varied population densities and healthcare access levels. These counties also represent a mix of socioeconomic conditions, with disparities in insurance coverage, income levels, and healthcare access (U.S. Census Bureau, 2025)

### 2.2. Data Sources

#### Wastewater Data

SARS-CoV-2 RNA concentration data were collected from six WWTPs from April 19, 2020, to July 1, 2021. RNA was quantified using RT-qPCR for samples collected before December 16, 2020, and RT-ddPCR for those collected thereafter. Wastewater samples were collected biweekly (Swift et al., 2023). We substituted the undetected SARS-CoV-2 concentrations with the limit of detection of 520 genome copies/L (Swift et al., 2023).

#### Socio-Demographic Data

The study incorporates socio-demographic and socioeconomic variables: age groups, gender, race/ethnicity, insurance coverage, and socioeconomic indicators sourced from the U.S. Census Bureau (U.S. Census Bureau, 2025). Age groups are categorized into four ranges: 0–19, 20–44, 45–64, and 65+ years, for an age-stratified analysis. Gender is considered as male and female, ensuring representation of potential differences in health risks. Race and ethnicity are classified into four groups: white, black or african american, hispanic, and other, to capture racial and ethnic backgrounds. Insurance coverage is categorized into medicare, medicaid, private, and uninsured, reflecting differences in healthcare access. Additionally, key socioeconomic indicators such as median household income, unemployment rate, labor force participation and social vulnerability index (SVI) are included. These variables allowed us to take into account the broader social determinants that influence disease susceptibility and healthcare access. All the socio-demographic data is obtained at ZIP code and aggregated at WWTPs (in appendix Table A.1).

#### Hospitalization Data

Daily COVID-19 hospitalization data were collected from medical claims records, which were obtained from the South Carolina Revenue and Fiscal Affairs Office (South Carolina Revenue and Fiscal Affairs Office, 2025). These data served as the outcome variable for model training and evaluation.

### 2.3. Data Preprocessing

This retrospective observational study aimed to predict COVID-19 hospitalizations using SARS-CoV-2 RNA concentrations in wastewater and socio-demographic variables. We explored the relationship between RNA levels and hospitalization trends at both broader (WWTP) and granular (ZIP code) scales. We analyzed data from April 19, 2020, to February 2, 2021, a period selected to capture the peak surge in hospitalizations. A sharp increase in cases was observed from December 2020, coinciding with the emergence of the Alpha variant in the USA(Centers for Disease Control and Prevention, 2023). We split the dataset into training and testing sets using a 90:10 ratio for both WWTP and ZIP code level analyses. Hospitalization data was smoothed using a 7-day moving average to reduce the noise inherent in daily reporting. For the first six days, the moving average was computed using available preceding values until the full 7-day window was established. On the other hand, wastewater samples were collected only twice a week. Historically, wastewater data has been very noisy. For this, we applied penalized spline regression with cubic B-spline basis functions to smooth the data and interpolate for the unsampled days. Before smoothing, we replaced zero RNA copies with 520 genome copies/L, as per the literature. By combining interpolated and smoothed values, we get continuous RNA concentration over time. ZIP codes that overlapped partially with a WWTP were included in full to ensure consistency in geographic coverage.

A population-weighted distribution approach was employed to estimate viral RNA concentrations at the ZIP code level. This method allocated RNA concentrations measured at WWTPs to individual ZIP codes based on their proportion of the total population served by the WWTP. For a given ZIP code i, the estimated RNA concentration was calculated as:

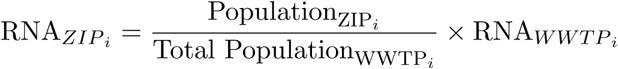

Here, RNA_ZIP*i*_ and Population_ZIP*i*_ denotes the estimated RNA concentration and the population of ZIP code *i*, Total Population_WWTP*i*_ is the total population served by the WWTP covering ZIP code *i*. RNA_WWTP_*_i_* denotes the measured RNA concentration at that WWTP. This approach ensured that ZIP codes with larger populations were assigned a proportionally greater share of the RNA concentration.

### 2.4. Analysis

We utilized SARS-CoV-2 RNA concentrations in wastewater on day t to predict COVID-19 hospitalizations at future time points (t + 7, t+14, and t+21 days) at the WWTP and ZIP code levels. This is equivalent to assuming a 7-day, 14-day, and 21-day lag between wastewater and hospitalization. Two predictive models were trained, Poisson regression and random forest, incorporating both RNA copies and socio-demographic variables. Poisson regression was employed to ensure that predicted hospitalization counts remained realistic and non-negative, while the random forest model captured potential nonlinear interactions among predictors. Model performance was evaluated using the Median Percentage Agreement (MPA) over a 28-day prediction horizon.

**Poisson Regression Model:** Poisson regression was implemented to model count data, specifically the number of hospitalizations. The model’s equation is:

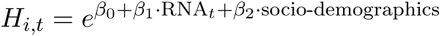

Where *H_i,t_* represents the expected number of hospitalizations for region *i* at time *t*, RNA*_t_* denotes the SARS-CoV-2 RNA concentrations at time *t*, and socio-demographics includes socio-demographic variables such as age, sex, race, and income etc.

**Random Forest Model:** Random Forest models, an ensemble learning method. The model leverages multiple decision trees, where each tree predicts hospitalizations independently, and the final prediction is the average of all trees:

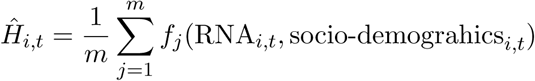

Where *f_j_*(*·*): The prediction from the *j*-th tree in the ensemble, *m*: Total number of decision trees.

**Accuracy of the Models** Model accuracy was evaluated using the Median Percentage Agreement (MPA) metric. The average percentage agreement for region *i* is defined as:

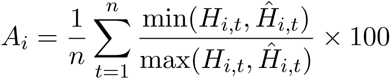

Where:

- *H_i,t_* is the observed number of hospitalizations for region *i* in week *t*,
- *H*^^^*_i,t_* is the predicted number of hospitalizations for region *i* in week *t*.

Daily predictions and observations were first aggregated by week (i.e., the sum of all daily hospitalizations within each 7-day period). The weekly totals were then compared using the equation above to calculate the average percentage agreement for each region. The MPA was computed by taking the median value of *A_i_* values across all areas.

## 3. Results

The predictive framework demonstrated reliable performance in forecasting COVID-19 hospitalizations using SARS-CoV-2 RNA concentrations from wastewater. We evaluated models with 7, 14, and 21 days ahead to account for the delay between viral shedding and hospitalization. This corresponds to the prediction of hospitalizations 7 days, 14 days, and 21 days after wastewater collection. Each model’s performance was evaluated over a 28-day test period. We explored multiple lag days, as viral shedding in wastewater precedes hospitalizations by days to weeks. The results discussed here represent the aggregated performance of models across all WWTPs (Table 1) and across all ZIP codes (Table 2).For both WWTP- and ZIP code–level predictions, we used the default settings of Random Forest and Poisson regression models from the scikit-learn library, as these configurations yielded reliable performance.

**Table 1:** Median percentage agreement of predicted hospitalizations across all wastewater treatment plants (WWTPs), utilizing COVID-19 RNA concentrations and socio-demographic variables with 7, 14, and 21 days ahead. Each model’s performance was evaluated over a 28-day test period. The highest accuracy is highlighted.

| Model | 7-Day Ahead | 14-Day Ahead | 21-Day Ahead |
| --- | --- | --- | --- |
| Poisson Regression | 77.44% (IQR: 76.46–79.37%) | 82.67% (IQR: 74.74–87.84%) | 74.69 % (IQR: 71.41–78.93%) |
| Random Forest | 86.44% (IQR: 79.25–89.85%) | **91.16% (IQR: 86.49–91.84%)** | 90.86 % (IQR: 88.73–90.86%) |

**Table 2:** Median percent agreement of predicted hospitalizations across all ZIP codes, using COVID-19 RNA concentrations in wastewater for 7-day, 14-day, and 21-days ahead. Each model’s performance was evaluated over a 28-day test period. The highest accuracy is highlighted.

| Model | 7-Day Ahead | 14-Day Ahead | 21-Day Ahead |
| --- | --- | --- | --- |
| Poisson Regression | 73.69% (IQR: 64.11–82.3%) | 74.31% (IQR: 63.74–79.98%) | 67.05% (IQR: 62.11–75.5%) |
| Random Forest | 75.15 (IQR: 66.3–84.93%) | **78.12% (IQR: 67.99–84.53%)** | 77.49% (IQR: 66.1–84.54%) |

### 3.1. WWTP-Level Analysis

At the WWTP level, the random forest consistently outperformed poison regression (Table 1). For the 7-day ahead, it achieved a median MPA of 86.44% (IQR: 79.25–89.85%). Random forests performance peaked for the 14-day horizon, reaching a median MPA of 91.16% (IQR: 86.49–91.84%). At 21-day predictions both the models exhibited lower accuracy. The detailed result is available in Table A.2 The results of the 14-day horizon random forest analysis are presented in Figure 1, and other results are available in the Figure A.1 and Figure A.2.

**Figure 1:**
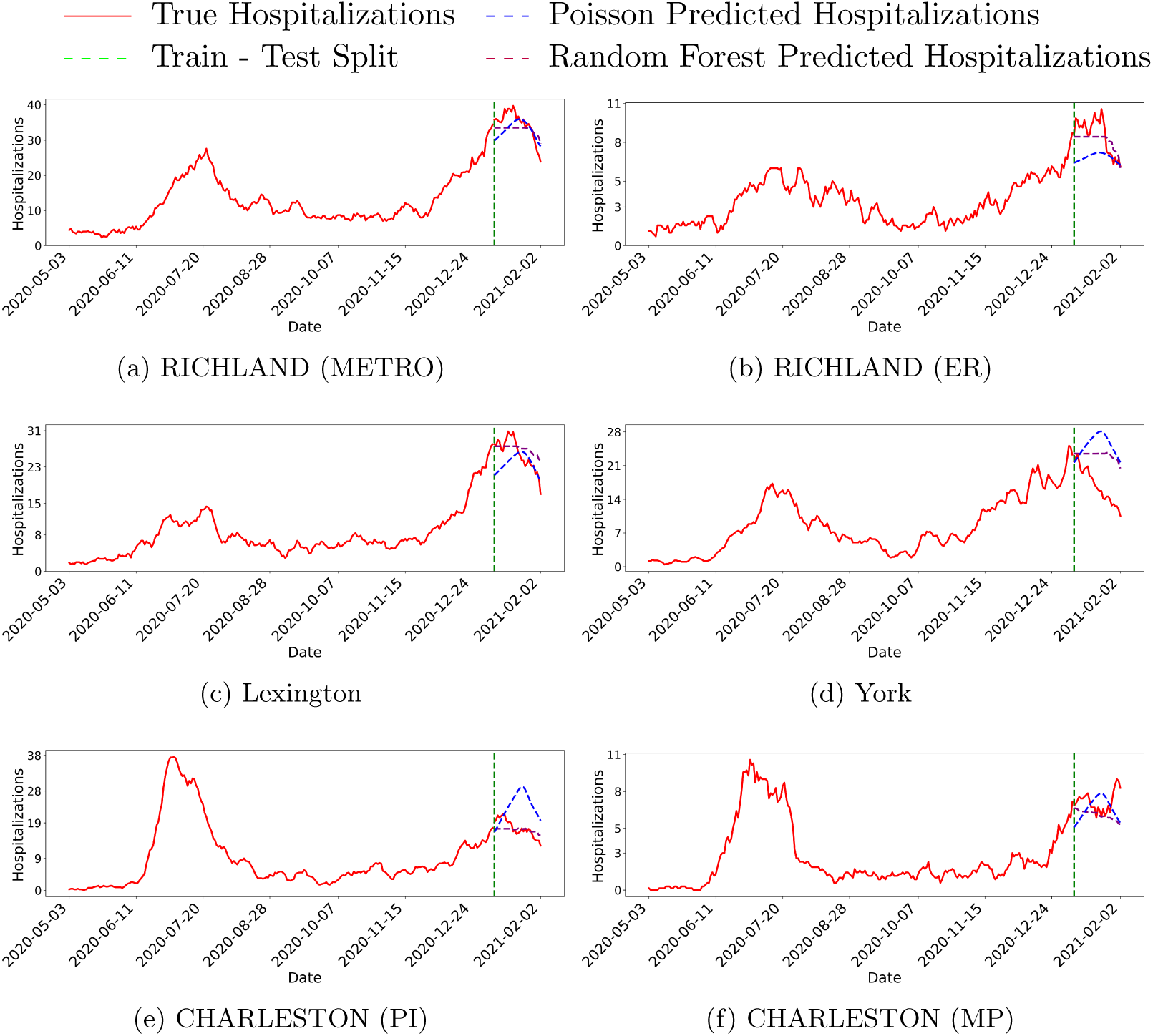
Predicted and true COVID-19 hospitalizations across wastewater treatment plants (WWTPs) using 14-day ahead. The purple line represents predictions from the random forest model, the blue line shows predictions from the Poisson regression model, and the red line indicates true hospitalizations. The dashed green line marks the point dividing the training and testing periods. 7-day and 21-day is available in appendix (Figure A.1 and Figure A.2)

### 3.2. ZIP Code-Level Analysis

At the ZIP code level, predictions exhibited greater variability due to the finer geographic granularity and inherent heterogeneity in population and data coverage. We utilized Poisson regression, which restricts the prediction to become negative. This was especially crucial for ZIP codes with small populations, low RNA concentrations, and sparse hospitalization data. In such cases, traditional models risked producing implausible negative values for hospitalizations, reducing the reliability of predictions. Random forest outperformed the Poisson regression model for all prediction horizons.

Both models peaked at 14 day ahead for the ZIP codes like the WWTPs. The random forest model achieved a median PA of 78.12% (IQR: 67.99–84.53%) and poison regression 74.31% (IQR: 63.74–79.98%) for the 14-day horizon. All associated figures and other results are available in the appendix (Table B.3).

## 4. Discussion

Accurate and timely forecasting of hospitalizations for COVID-19 remains critical for public health systems to allocate resources effectively (Assefa et al., 2022). WBE provides a state-of-the-art approach by monitoring community-level viral shedding, capturing data from symptomatic, asymptomatic, and untested individuals (Hillary et al., 2020; Hart and Halden, 2020). Our study demonstrates how WBE, combined with machine learning models, can bridge gaps in traditional surveillance. We aimed to develop a framework for predicting COVID-19 hospitalizations at the WWTP and ZIP code levels from SARS-CoV-2 RNA concentration in wastewater. By utilizing SARS-CoV-2 RNA concentrations and socio-demographic data, the model functions as an early warning system. This method enhances conventional health surveillance by providing finer geographic resolution.

Among the methods evaluated, the random forest model consistently emerged as the most reliable method for predicting hospitalizations at the WWTP level. The model achieved median MPAs of 86.44%, 91.16%, and 90.61% for the 7-, 14-day and 21-days ahead, respectively (Table 1). These results demonstrate the model’s ability to capture key trends in hospitalization from wastewater signals. In contrast, Poisson regression models achieved lower performance, particularly as the forecast window extended. At the ZIP code level, the predictions were generally lower due to the finer spatial resolution and population heterogeneity (Table 2). Predictions made 14 days ahead achieved the highest accuracy, which is consistent with studies showing an approximate 2-week lag between infection and hospital admission (Li et al., 2023; Galani et al., 2022; Hill et al., 2023).

The findings of this study have significant implications for public health planning and response at granular level. By integrating wastewater surveillance data with ML algorithms, this framework lays the groundwork for real-time prediction and monitoring. Additionally, the insights enable targeted resource allocation, allowing hospitals to make short time staffing and procure essential supplies in advance based on tailored local predictions. These predictions also allow local public health officials to prioritize testing campaigns, and deploying mobile health clinics [ref Tanvir, Sakhawat, and Fatih papers here]. The scalability of this framework to a granular level suggests potential applicability beyond COVID-19. However, it’s important to note that different infections display unique viral shedding patterns.

Despite its predictive strengths, this study has some limitations. First, the retrospective nature of the analysis restricts its use for real-time prediction, highlighting the necessity of adapting the framework for proactive forecasting. Second, our research focused specifically on South Carolina and a limited subset of wastewater treatment plants (WWTPs), which may affect the generalizability of the findings. While this study demonstrates the utility of an ML-based approach for granular hospitalization forecasting, these models should be retrained when adaptating to other geographic regions, time periods, or diseases. Lastly, one critical factor to consider in WBE is the impact of septic tanks. Since septic systems are not connected to a centralized sampling point, monitoring viral transmission in regions lacking centralized systems becomes more complex. However, this limitation is counterbalanced by the low cost and straightforward implementation of WBE. This makes it particularly beneficial in developing countries with limited surveillance resources. Future research should focus on real-time forecasting while expanding the geographic scope to increase model reliability. Incorporating emerging variants, mobility patterns, and vaccination rates will improve adaptability to evolving health threats. Increasing the frequency of wastewater sampling could augment prediction accuracy, particularly at the ZIP code levels.

## 5. Conclusion

This study demonstrates the feasibility of using wastewater surveillance and machine learning to estimate COVID-19 hospitalizations. The random forest model outperformed Poisson regression, providing reliable predictions with RNA data and geographic variables. By integrating SARS-CoV-2 RNA concentrations with socio-demographic data, this framework improves disease monitoring and public health response. It enables targeted resource allocation, such as optimizing hospital capacity and vaccination efforts. Its scalability allows application to other infectious diseases and public health crises. Machine learning helps address data gaps, ensuring reliable estimates even in areas with limited direct data. Though retrospective, this framework supports real-time monitoring with potential improvements from updated data and evolving pandemic dynamics. Expanding our framework to include environmental and behavioral factors could enhance its predictive power. This study advances wastewater-based epidemiology, offering a scalable and cost-effective tool for public health. Future research should extend its use across regions and health events to strengthen preparedness.

## Author Statement

LR, CM, and RSN conceptualized and supervised this study. NT and MMC conducted all data analyses and visualizations with contributions from LR, CM. LR, CM, NT, and MMC developed the study methodology. SSelf, MI, KCV, SSellers, and RSN collected and curated the wastewater data. NT and MMC wrote the original draft, with contributions from LR. LR performed project administration and acquired funding. All authors interpreted the study findings, reviewed and edited the manuscript, had access to all data, approved the final version, agreed to be accountable for all aspects of the work, and had final responsibility for the decision to submit for publication.

## Ethics Statement

Ethical review for this study was obtained by the Institutional Review Board of Clemson University (#2020-0150). No consent was needed for this study; retrospective data is based on wastewater and medical claims and were de-identified to study investigators.

## Data Availability

All data produced in the present study are available upon reasonable request to the authors

## Acknowledgments

This project has been funded by the Center for Forecasting and Outbreak Analytics of the Centers for Disease Control and Prevention (CDC) under Award no. NU38FT000011 and the National Library of Medicine of the National Institutes of Health (NIH) under Award no. R01LM014193. The content and decision to publish is solely based on the authors of this study and does not necessarily represent the official views of the CDC or NIH. The funders had no role in the study design, data collection and analysis, decision to publish, or preparation of this manuscript.

## Appendix

### A. WWTP

**Table A.1:** Socio-Demographic Characteristics of WWTP Service Areas. Population estimates were based on 2019 U.S. Census ZIP code level data. ZIP codes partially within a WWTP boundary were included in full. (U.S. Census Bureau, 2025)

| WWTP Area | Richland (Metro) | Richland (ER) | Lexington | York | Charleston (PI*) | Charleston (MP*) |
| --- | --- | --- | --- | --- | --- | --- |
| **Sex** |  |  |  |  |  |  |
| Male | 182888 (49%) | 41813 (47%) | 114108 (49%) | 128368 (49%) | 98083 (47%) | 47402 (49%) |
| Female | 188787 (51%) | 46461 (53%) | 117980 (51%) | 136152 (51%) | 109533 (53%) | 50103 (51%) |
| **Age Group** |  |  |  |  |  |  |
| 0-19 years | 95101 (26%) | 21714 (25%) | 59554 (26%) | 70833 (27%) | 40293 (19%) | 23249 (24%) |
| 20-44 years | 141674 (38%) | 30156 (34%) | 75252 (32%) | 84641 (32%) | 80089 (39%) | 29383 (30%) |
| 45-64 years | 83890 (23%) | 21362 (24%) | 61923 (27%) | 71100 (27%) | 52138 (25%) | 26995 (28%) |
| ≥ 65 years | 51010 (14%) | 15042 (17%) | 35359 (15%) | 37946 (14%) | 35096 (17%) | 17878 (18%) |
| **Race or Ethnicity** |  |  |  |  |  |  |
| Hispanic | 18706 (5%) | 5328 (6%) | 15611 (7%) | 16250 (6%) | 7696 (4%) | 2165 (2%) |
| White | 176720 (48%) | 36574 (41%) | 177602 (77%) | 190438 (72%) | 151656 (73%) | 84708 (87%) |
| Black | 161656 (43%) | 42864 (49%) | 32436 (14%) | 50346 (19%) | 44279 (21%) | 7915 (8%) |
| Other | 33299 (9%) | 8836 (10%) | 22050 (10%) | 23736 (9%) | 11681 (6%) | 4882 (5%) |
| **Socioeconomic** |  |  |  |  |  |  |
| SVI | 0.438 | 0.533 | 0.481 | 0.331 | 0.290 | 0.181 |
| Median Income | 691828 | 174093 | 611435 | 471923 | 692806 | 282019 |
| Employed Labor | 180853 (49%) | 42157 (48%) | 112030 (48%) | 131042 (50%) | 114350 (55%) | 49440 (51%) |
| Unemployed Labor | 11632 (3%) | 2572 (3%) | 5471 (2%) | 6619 (3%) | 3818 (2%) | 1997 (2%) |
| **Healthcare Access** |  |  |  |  |  |  |
| Total Hospital | 15 | 1 | 4 | 3 | 4 | 3 |
| Private | 180243 (48%) | 40439 (46%) | 121450 (52%) | 161626 (61%) | 130119 (63%) | 63159 (65%) |
| Medicare | 14227 (4%) | 4325 (5%) | 10935 (5%) | 12783 (5%) | 12290 (6%) | 5766 (6%) |
| Medicaid | 50198 (14%) | 12419 (14%) | 30269 (13%) | 27792 (11%) | 13687 (7%) | 3799 (4%) |
| Uninsured | 32593 (9%) | 8565 (10%) | 22655 (10%) | 21791 (8%) | 14891 (7%) | 3947 (4%) |

PI\* = Plum Island
MP\* = Mount Pleasant

**Figure A.1:**
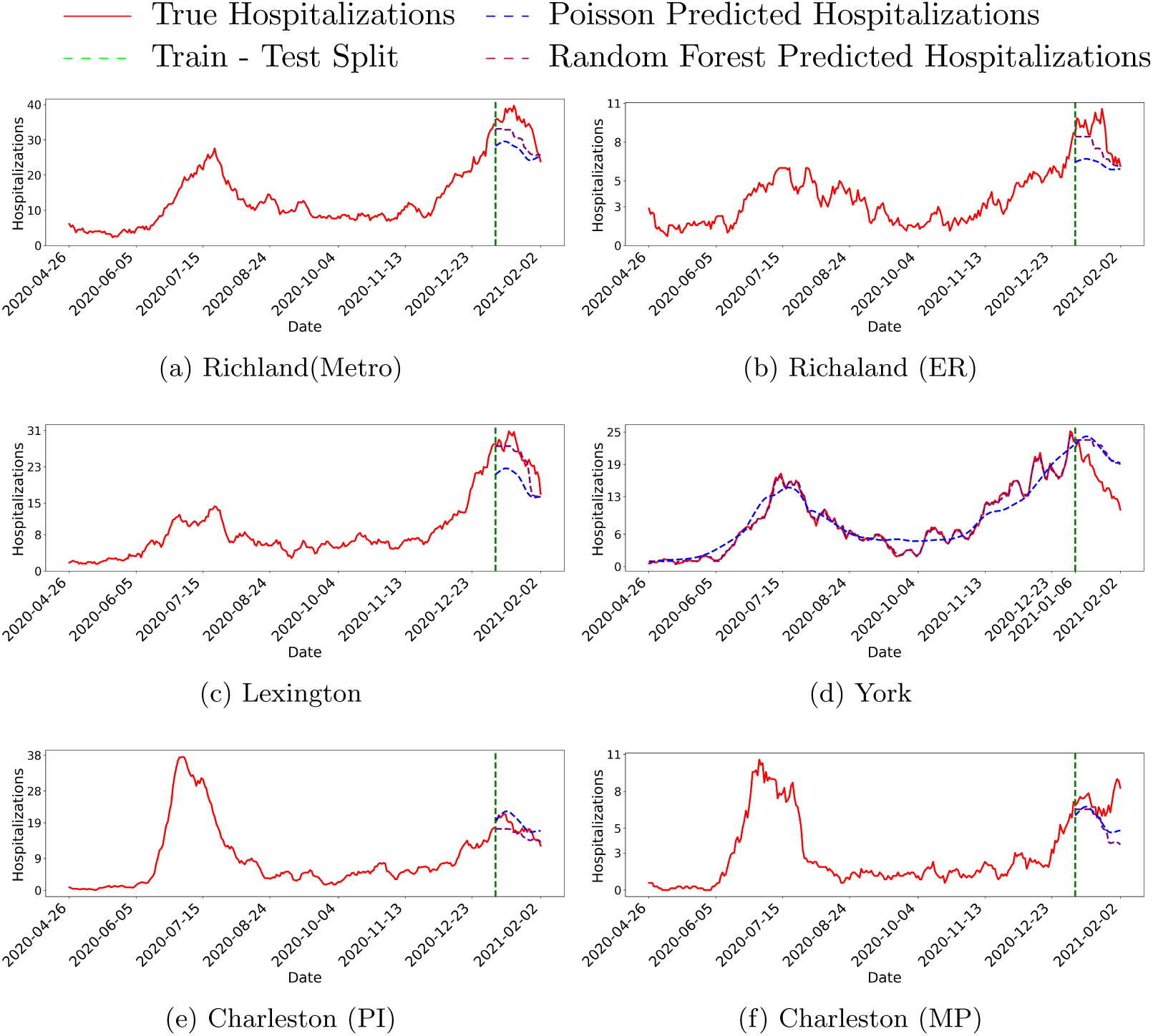
Predicted and true COVID-19 hospitalizations across wastewater treatment plants (WWTPs) using 7-day ahead. The purple line represents predictions from the random forest model, the blue line shows predictions from the Poisson regression model, and the red line indicates true hospitalizations. The dashed green line marks the point dividing the training and testing periods.

**Figure A.2:**
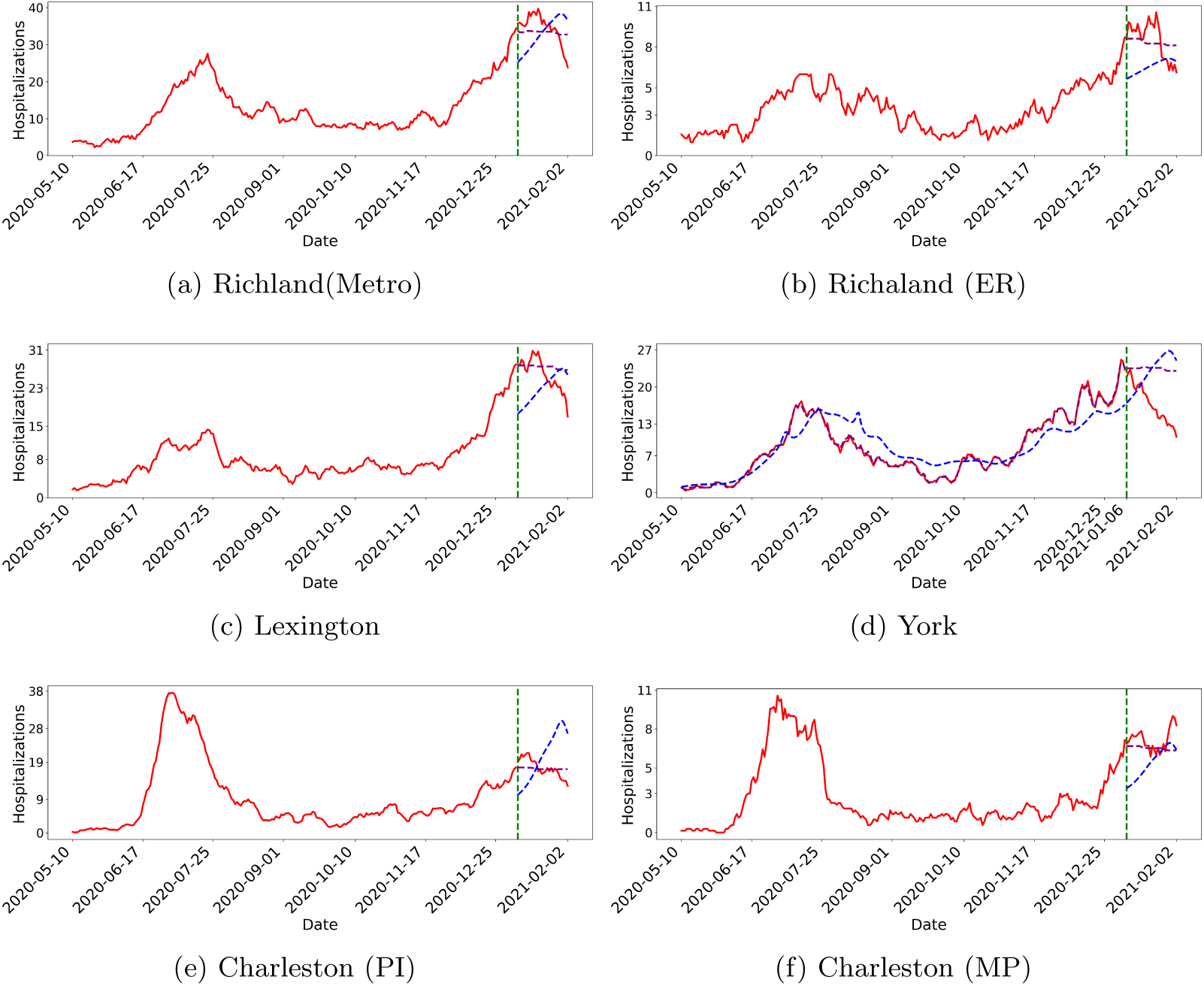
Predicted and true COVID-19 hospitalizations across wastewater treatment plants (WWTPs) using 21-day ahead. The purple line represents predictions from the random forest model, the blue line shows predictions from the Poisson regression model, ahospitalisation indicates true hospitalizations. The dashed green line marks the point dividing the training and testing periods.

**Table A.2:** Average Percentage Agreement over the prediction period Between Predicted and Observed COVID-19 Hospitalizations, Stratified by Modeling Approach, Days Ahead, and Wastewater Treatment Plant (WWTP) Service Area

| WWTP | Model | Days Ahead | Average Percent Agreement |
| --- | --- | --- | --- |
| Richland (Metro) | Poisson Regression | 5 | 74.48 |
| Richland (Metro) | Poisson Regression | 7 | 78.29 |
| Richland (Metro) | Poisson Regression | 14 | 92.68 |
| Richland (Metro) | Poisson Regression | 21 | 83.04 |
| Richland (Metro) | Random Forest | 5 | 82.01 |
| Richland (Metro) | Random Forest | 7 | 86.13 |
| Richland (Metro) | Random Forest | 14 | 91.87 |
| Richland (Metro) | Random Forest | 21 | 91.11 |
| Richland (ER) | Poisson Regression | 5 | 72.01 |
| Richland (ER) | Poisson Regression | 7 | 74.58 |
| Richland (ER) | Poisson Regression | 14 | 80.86 |
| Richland (ER) | Poisson Regression | 21 | 74.88 |
| Richland (ER) | Random Forest | 5 | 82.21 |
| Richland (ER) | Random Forest | 7 | 86.75 |
| Richland (ER) | Random Forest | 14 | 90.57 |
| Richland (ER) | Random Forest | 21 | 88.21 |

| WWTP | Model | Days Ahead) | Average Percent Agreement |
| --- | --- | --- | --- |
| Lexington | Poisson Regression | 5 | 72.94 |
| Lexington | Poisson Regression | 7 | 76.42 |
| Lexington | Poisson Regression | 14 | 88.96 |
| Lexington | Poisson Regression | 21 | 80.28 |
| Lexington | Random Forest | 5 | 86.51 |
| Lexington | Random Forest | 7 | 91.83 |
| Lexington | Random Forest | 14 | 91.75 |
| Lexington | Random Forest | 21 | 90.57 |
| York | Poisson Regression | 5 | 79.39 |
| York | Poisson Regression | 7 | 76.58 |
| York | Poisson Regression | 14 | 67.93 |
| York | Poisson Regression | 21 | 70.38 |
| York | Random Forest | 5 | 78.10 |
| York | Random Forest | 7 | 76.96 |
| York | Random Forest | 14 | 73.10 |
| York | Random Forest | 21 | 72.38 |
| Charleston (PI) | Poisson Regression | 5 | 95.21 |
| Charleston (PI) | Poisson Regression | 7 | 91.06 |
| Charleston (PI) | Poisson Regression | 14 | 72.70 |
| Charleston (PI) | Poisson Regression | 21 | 68.68 |
| Charleston (PI) | Random Forest | 5 | 88.59 |
| Charleston (PI) | Random Forest | 7 | 90.88 |
| Charleston (PI) | Random Forest | 14 | 91.95 |
| Charleston (PI) | Random Forest | 21 | 91.21 |
| Charleston (MP) | Poisson Regression | 5 | 75.24 |
| Charleston (MP) | Poisson Regression | 7 | 79.73 |
| Charleston (MP) | Poisson Regression | 14 | 84.48 |
| Charleston (MP) | Poisson Regression | 21 | 74.50 |
| Charleston (MP) | Random Forest | 5 | 74.77 |
| Charleston (MP) | Random Forest | 7 | 76.01 |
| Charleston (MP) | Random Forest | 14 | 85.13 |
| Charleston (MP) | Random Forest | 21 | 90.38 |

### B. ZIP code

**Table B.3:** Average Percentage Agreement in the Prediction Period Between Predicted and Observed COVID-19 Hospitalizations, Stratified by Modeling Approach, Days Ahead, and ZIP code

| ZIP code | Model | Days Ahead | Average Percentage Agreement |
| --- | --- | --- | --- |
| 29016 | Poisson Regression | 5 | 83.50 |
| 29016 | Poisson Regression | 7 | 86.32 |
| 29016 | Poisson Regression | 14 | 77.90 |
| 29016 | Poisson Regression | 21 | 75.46 |
| 29016 | Random Forest | 5 | 84.26 |
| 29016 | Random Forest | 7 | 82.25 |
| 29016 | Random Forest | 14 | 79.05 |
| 29016 | Random Forest | 21 | 78.34 |
| 29033 | Poisson Regression | 5 | 77.33 |
| 29033 | Poisson Regression | 7 | 80.89 |
| 29033 | Poisson Regression | 14 | 80.18 |
| 29033 | Poisson Regression | 21 | 88.08 |
| 29033 | Random Forest | 5 | 88.33 |
| 29033 | Random Forest | 7 | 85.52 |
| 29033 | Random Forest | 14 | 83.77 |
| 29033 | Random Forest | 21 | 85.40 |
| 29053 | Poisson Regression | 5 | 60.22 |
| 29053 | Poisson Regression | 7 | 61.87 |
| 29053 | Poisson Regression | 14 | 78.26 |
| 29053 | Poisson Regression | 21 | 68.54 |
| 29053 | Random Forest | 5 | 74.97 |
| 29053 | Random Forest | 7 | 77.68 |
| 29053 | Random Forest | 14 | 87.80 |
| 29053 | Random Forest | 21 | 86.29 |
| 29054 | Poisson Regression | 5 | 61.52 |
| 29054 | Poisson Regression | 7 | 64.72 |
| 29054 | Poisson Regression | 14 | 68.18 |
| 29054 | Poisson Regression | 21 | 62.20 |
| 29054 | Random Forest | 5 | 67.41 |
| 29054 | Random Forest | 7 | 63.52 |
| 29054 | Random Forest | 14 | 67.01 |
| 29054 | Random Forest | 21 | 62.64 |
| 29063 | Poisson Regression | 5 | 85.28 |
| 29063 | Poisson Regression | 7 | 87.07 |
| 29063 | Poisson Regression | 14 | 82.44 |
| 29063 | Poisson Regression | 21 | 77.99 |
| 29063 | Random Forest | 5 | 85.25 |
| 29063 | Random Forest | 7 | 88.83 |
| 29063 | Random Forest | 14 | 91.69 |
| 29063 | Random Forest | 21 | 92.28 |
| 29070 | Poisson Regression | 5 | 68.83 |
| 29070 | Poisson Regression | 7 | 75.52 |
| 29070 | Poisson Regression | 14 | 85.35 |
| 29070 | Poisson Regression | 21 | 75.54 |
| 29070 | Random Forest | 5 | 84.12 |
| 29070 | Random Forest | 7 | 87.01 |
| 29070 | Random Forest | 14 | 85.38 |
| 29070 | Random Forest | 21 | 83.34 |
| 29072 | Poisson Regression | 5 | 86.77 |
| 29072 | Poisson Regression | 7 | 91.99 |
| 29072 | Poisson Regression | 14 | 86.05 |
| 29072 | Poisson Regression | 21 | 79.44 |
| 29072 | Random Forest | 5 | 90.03 |
| 29072 | Random Forest | 7 | 91.45 |
| 29072 | Random Forest | 14 | 84.03 |
| 29072 | Random Forest | 21 | 83.16 |
| 29073 | Poisson Regression | 5 | 73.95 |
| 29073 | Poisson Regression | 7 | 75.80 |
| 29073 | Poisson Regression | 14 | 74.94 |
| 29073 | Poisson Regression | 21 | 68.03 |
| 29073 | Random Forest | 5 | 80.61 |
| 29073 | Random Forest | 7 | 78.68 |
| 29073 | Random Forest | 14 | 76.33 |
| 29073 | Random Forest | 21 | 75.99 |
| 29160 | Poisson Regression | 5 | 62.81 |
| 29160 | Poisson Regression | 7 | 59.33 |
| 29160 | Poisson Regression | 14 | 59.77 |
| 29160 | Poisson Regression | 21 | 66.16 |
| 29160 | Random Forest | 5 | 76.80 |
| 29160 | Random Forest | 7 | 73.13 |
| 29160 | Random Forest | 14 | 72.11 |
| 29160 | Random Forest | 21 | 71.88 |
| 29169 | Poisson Regression | 5 | 59.48 |
| 29169 | Poisson Regression | 7 | 62.61 |
| 29169 | Poisson Regression | 14 | 62.24 |
| 29169 | Poisson Regression | 21 | 50.39 |
| 29169 | Random Forest | 5 | 64.19 |
| 29169 | Random Forest | 7 | 66.26 |
| 29169 | Random Forest | 14 | 78.12 |
| 29169 | Random Forest | 21 | 79.98 |
| 29170 | Poisson Regression | 5 | 65.82 |
| 29170 | Poisson Regression | 7 | 68.85 |
| 29170 | Poisson Regression | 14 | 81.19 |
| 29170 | Poisson Regression | 21 | 72.87 |
| 29170 | Random Forest | 5 | 70.44 |
| 29170 | Random Forest | 7 | 74.41 |
| 29170 | Random Forest | 14 | 81.75 |
| 29170 | Random Forest | 21 | 78.96 |
| 29172 | Poisson Regression | 5 | 69.27 |
| 29172 | Poisson Regression | 7 | 69.48 |
| 29172 | Poisson Regression | 14 | 74.31 |
| 29172 | Poisson Regression | 21 | 66.55 |
| 29172 | Random Forest | 5 | 58.34 |
| 29172 | Random Forest | 7 | 59.90 |
| 29172 | Random Forest | 14 | 68.97 |
| 29172 | Random Forest | 21 | 66.62 |
| 29201 | Poisson Regression | 5 | 60.80 |
| 29201 | Poisson Regression | 7 | 62.89 |
| 29201 | Poisson Regression | 14 | 68.21 |
| 29201 | Poisson Regression | 21 | 70.26 |
| 29201 | Random Forest | 5 | 64.84 |
| 29201 | Random Forest | 7 | 65.98 |
| 29201 | Random Forest | 14 | 71.50 |
| 29201 | Random Forest | 21 | 73.36 |
| 29203 | Poisson Regression | 5 | 72.37 |
| 29203 | Poisson Regression | 7 | 77.52 |
| 29203 | Poisson Regression | 14 | 88.26 |
| 29203 | Poisson Regression | 21 | 80.53 |
| 29203 | Random Forest | 5 | 89.81 |
| 29203 | Random Forest | 7 | 86.00 |
| 29203 | Random Forest | 14 | 92.49 |
| 29203 | Random Forest | 21 | 90.86 |
| 29204 | Poisson Regression | 5 | 52.30 |
| 29204 | Poisson Regression | 7 | 53.15 |
| 29204 | Poisson Regression | 14 | 56.00 |
| 29204 | Poisson Regression | 21 | 57.61 |
| 29204 | Random Forest | 5 | 74.39 |
| 29204 | Random Forest | 7 | 82.95 |
| 29204 | Random Forest | 14 | 87.60 |
| 29204 | Random Forest | 21 | 85.91 |
| 29205 | Poisson Regression | 5 | 63.53 |
| 29205 | Poisson Regression | 7 | 65.83 |
| 29205 | Poisson Regression | 14 | 68.79 |
| 29205 | Poisson Regression | 21 | 63.42 |
| 29205 | Random Forest | 5 | 71.79 |
| 29205 | Random Forest | 7 | 74.93 |
| 29205 | Random Forest | 14 | 64.09 |
| 29205 | Random Forest | 21 | 65.55 |
| 29206 | Poisson Regression | 5 | 71.11 |
| 29206 | Poisson Regression | 7 | 72.60 |
| 29206 | Poisson Regression | 14 | 72.90 |
| 29206 | Poisson Regression | 21 | 66.27 |
| 29206 | Random Forest | 5 | 80.96 |
| 29206 | Random Forest | 7 | 78.32 |
| 29206 | Random Forest | 14 | 76.27 |
| 29206 | Random Forest | 21 | 75.54 |
| 29209 | Poisson Regression | 5 | 81.86 |
| 29209 | Poisson Regression | 7 | 83.05 |
| 29209 | Poisson Regression | 14 | 80.38 |
| 29209 | Poisson Regression | 21 | 70.30 |
| 29209 | Random Forest | 5 | 81.56 |
| 29209 | Random Forest | 7 | 80.03 |
| 29209 | Random Forest | 14 | 80.47 |
| 29209 | Random Forest | 21 | 72.27 |
| 29210 | Poisson Regression | 5 | 64.21 |
| 29210 | Poisson Regression | 7 | 65.84 |
| 29210 | Poisson Regression | 14 | 77.16 |
| 29210 | Poisson Regression | 21 | 82.35 |
| 29210 | Random Forest | 5 | 66.03 |
| 29210 | Random Forest | 7 | 62.50 |
| 29210 | Random Forest | 14 | 83.62 |
| 29210 | Random Forest | 21 | 72.50 |
| 29212 | Poisson Regression | 5 | 83.47 |
| 29212 | Poisson Regression | 7 | 89.48 |
| 29212 | Poisson Regression | 14 | 87.93 |
| 29212 | Poisson Regression | 21 | 76.09 |
| 29212 | Random Forest | 5 | 85.60 |
| 29212 | Random Forest | 7 | 87.16 |
| 29212 | Random Forest | 14 | 89.04 |
| 29212 | Random Forest | 21 | 88.15 |
| 29223 | Poisson Regression | 5 | 83.52 |
| 29223 | Poisson Regression | 7 | 87.01 |
| 29223 | Poisson Regression | 14 | 83.07 |
| 29223 | Poisson Regression | 21 | 76.43 |
| 29223 | Random Forest | 5 | 83.87 |
| 29223 | Random Forest | 7 | 86.85 |
| 29223 | Random Forest | 14 | 85.01 |
| 29223 | Random Forest | 21 | 87.55 |
| 29401 | Poisson Regression | 5 | 49.02 |
| 29401 | Poisson Regression | 7 | 50.86 |
| 29401 | Poisson Regression | 14 | 49.79 |
| 29401 | Poisson Regression | 21 | 44.24 |
| 29401 | Random Forest | 5 | 31.22 |
| 29401 | Random Forest | 7 | 30.69 |
| 29401 | Random Forest | 14 | 31.79 |
| 29401 | Random Forest | 21 | 48.27 |
| 29403 | Poisson Regression | 5 | 58.59 |
| 29403 | Poisson Regression | 7 | 60.55 |
| 29403 | Poisson Regression | 14 | 75.95 |
| 29403 | Poisson Regression | 21 | 63.22 |
| 29403 | Random Forest | 5 | 46.14 |
| 29403 | Random Forest | 7 | 48.55 |
| 29403 | Random Forest | 14 | 52.64 |
| 29403 | Random Forest | 21 | 51.16 |
| 29407 | Poisson Regression | 5 | 87.53 |
| 29407 | Poisson Regression | 7 | 83.05 |
| 29407 | Poisson Regression | 14 | 66.16 |
| 29407 | Poisson Regression | 21 | 60.07 |
| 29407 | Random Forest | 5 | 85.83 |
| 29407 | Random Forest | 7 | 84.63 |
| 29407 | Random Forest | 14 | 82.36 |
| 29407 | Random Forest | 21 | 81.58 |
| 29412 | Poisson Regression | 5 | 74.01 |
| 29412 | Poisson Regression | 7 | 71.86 |
| 29412 | Poisson Regression | 14 | 63.86 |
| 29412 | Poisson Regression | 21 | 74.49 |
| 29412 | Random Forest | 5 | 74.23 |
| 29412 | Random Forest | 7 | 72.65 |
| 29412 | Random Forest | 14 | 76.39 |
| 29412 | Random Forest | 21 | 77.55 |
| 29414 | Poisson Regression | 5 | 81.24 |
| 29414 | Poisson Regression | 7 | 76.17 |
| 29414 | Poisson Regression | 14 | 65.68 |
| 29414 | Poisson Regression | 21 | 66.17 |
| 29414 | Random Forest | 5 | 83.17 |
| 29414 | Random Forest | 7 | 85.69 |
| 29414 | Random Forest | 14 | 87.20 |
| 29414 | Random Forest | 21 | 85.23 |
| 29429 | Poisson Regression | 5 | 58.60 |
| 29429 | Poisson Regression | 7 | 60.26 |
| 29429 | Poisson Regression | 14 | 52.38 |
| 29429 | Poisson Regression | 21 | 32.97 |
| 29429 | Random Forest | 5 | 21.96 |
| 29429 | Random Forest | 7 | 23.82 |
| 29429 | Random Forest | 14 | 28.78 |
| 29429 | Random Forest | 21 | 33.80 |
| 29449 | Poisson Regression | 5 | 83.64 |
| 29449 | Poisson Regression | 7 | 83.02 |
| 29449 | Poisson Regression | 14 | 66.26 |
| 29449 | Poisson Regression | 21 | 62.02 |
| 29449 | Random Forest | 5 | 83.69 |
| 29449 | Random Forest | 7 | 88.72 |
| 29449 | Random Forest | 14 | 79.00 |
| 29449 | Random Forest | 21 | 85.00 |
| 29455 | Poisson Regression | 5 | 74.62 |
| 29455 | Poisson Regression | 7 | 78.45 |
| 29455 | Poisson Regression | 14 | 75.10 |
| 29455 | Poisson Regression | 21 | 66.53 |
| 29455 | Random Forest | 5 | 63.21 |
| 29455 | Random Forest | 7 | 66.33 |
| 29455 | Random Forest | 14 | 65.99 |
| 29455 | Random Forest | 21 | 64.59 |
| 29464 | Poisson Regression | 5 | 91.45 |
| 29464 | Poisson Regression | 7 | 91.43 |
| 29464 | Poisson Regression | 14 | 66.78 |
| 29464 | Poisson Regression | 21 | 62.85 |
| 29464 | Random Forest | 5 | 85.23 |
| 29464 | Random Forest | 7 | 82.23 |
| 29464 | Random Forest | 14 | 85.13 |
| 29464 | Random Forest | 21 | 83.10 |
| 29466 | Poisson Regression | 5 | 72.18 |
| 29466 | Poisson Regression | 7 | 73.69 |
| 29466 | Poisson Regression | 14 | 80.96 |
| 29466 | Poisson Regression | 21 | 85.81 |
| 29466 | Random Forest | 5 | 71.12 |
| 29466 | Random Forest | 7 | 70.99 |
| 29466 | Random Forest | 14 | 84.61 |
| 29466 | Random Forest | 21 | 86.89 |
| 29470 | Poisson Regression | 5 | 66.10 |
| 29470 | Poisson Regression | 7 | 63.50 |
| 29470 | Poisson Regression | 14 | 63.62 |
| 29470 | Poisson Regression | 21 | 72.12 |
| 29470 | Random Forest | 5 | 78.95 |
| 29470 | Random Forest | 7 | 75.15 |
| 29470 | Random Forest | 14 | 77.62 |
| 29470 | Random Forest | 21 | 77.49 |
| 29492 | Poisson Regression | 5 | 81.86 |
| 29492 | Poisson Regression | 7 | 77.47 |
| 29492 | Poisson Regression | 14 | 61.02 |
| 29492 | Poisson Regression | 21 | 51.38 |
| 29492 | Random Forest | 5 | 70.71 |
| 29492 | Random Forest | 7 | 70.90 |
| 29492 | Random Forest | 14 | 61.07 |
| 29492 | Random Forest | 21 | 58.46 |
| 29708 | Poisson Regression | 5 | 72.98 |
| 29708 | Poisson Regression | 7 | 74.26 |
| 29708 | Poisson Regression | 14 | 79.78 |
| 29708 | Poisson Regression | 21 | 68.05 |
| 29708 | Random Forest | 5 | 63.80 |
| 29708 | Random Forest | 7 | 70.59 |
| 29708 | Random Forest | 14 | 71.47 |
| 29708 | Random Forest | 21 | 63.90 |
| 29710 | Poisson Regression | 5 | 91.72 |
| 29710 | Poisson Regression | 7 | 88.04 |
| 29710 | Poisson Regression | 14 | 78.02 |
| 29710 | Poisson Regression | 21 | 79.39 |
| 29710 | Random Forest | 5 | 86.12 |
| 29710 | Random Forest | 7 | 85.22 |
| 29710 | Random Forest | 14 | 84.45 |
| 29710 | Random Forest | 21 | 84.08 |
| 29715 | Poisson Regression | 5 | 57.02 |
| 29715 | Poisson Regression | 7 | 56.27 |
| 29715 | Poisson Regression | 14 | 55.50 |
| 29715 | Poisson Regression | 21 | 55.13 |
| 29715 | Random Forest | 5 | 62.33 |
| 29715 | Random Forest | 7 | 62.85 |
| 29715 | Random Forest | 14 | 66.24 |
| 29715 | Random Forest | 21 | 67.75 |
| 29730 | Poisson Regression | 5 | 69.57 |
| 29730 | Poisson Regression | 7 | 65.85 |
| 29730 | Poisson Regression | 14 | 52.09 |
| 29730 | Poisson Regression | 21 | 61.32 |
| 29730 | Random Forest | 5 | 68.89 |
| 29730 | Random Forest | 7 | 69.22 |
| 29730 | Random Forest | 14 | 66.81 |
| 29730 | Random Forest | 21 | 65.38 |
| 29732 | Poisson Regression | 5 | 81.22 |
| 29732 | Poisson Regression | 7 | 81.58 |
| 29732 | Poisson Regression | 14 | 74.96 |
| 29732 | Poisson Regression | 21 | 67.05 |
| 29732 | Random Forest | 5 | 81.59 |
| 29732 | Random Forest | 7 | 78.89 |
| 29732 | Random Forest | 14 | 72.82 |
| 29732 | Random Forest | 21 | 71.03 |
| 29745 | Poisson Regression | 5 | 68.83 |
| 29745 | Poisson Regression | 7 | 65.67 |
| 29745 | Poisson Regression | 14 | 50.86 |
| 29745 | Poisson Regression | 21 | 58.23 |
| 29745 | Random Forest | 5 | 63.21 |
| 29745 | Random Forest | 7 | 62.92 |
| 29745 | Random Forest | 14 | 60.25 |
| 29745 | Random Forest | 21 | 59.23 |

